# System-specific multimorbidity derived from prescribing data predicts colorectal cancer outcomes: a Scottish data-linkage study

**DOI:** 10.64898/2026.05.30.26354508

**Authors:** Karen N Barnett, Linda Williams, David Weller, Stewart W Mercer, Bruce Guthrie, Hester Ward, David H Brewster, Gill Hubbard, Christine Campbell

## Abstract

Multimorbidity, the co-existence of two or more long-term conditions, is up to three times more prevalent among people with cancer than in the general population and is associated with poorer survival, particularly for cancers with a more favourable prognosis such as colorectal cancer. In Scotland, multimorbidity is the norm among older adults, emerges earlier in socioeconomically deprived populations, and may contribute to comparatively low cancer survival rates. Despite this, the influence of multimorbidity on the colorectal cancer pathway remains poorly understood.

We conducted a Scottish data-linkage study of adults diagnosed with colorectal cancer between 2010 and 2014, linking the Scottish Cancer Registry to national prescribing, hospital admissions, death registration, and bowel screening datasets. Prescribing data were used to derive overall and system-specific comorbidity measures as a proxy for multimorbidity and active disease burden. Associations with stage at diagnosis, treatment, survival, and screening uptake were examined using logistic regression and Cox proportional hazards models adjusted for demographic and clinical covariates.

Among 19,043 patients, 87% had at least one prescribing-based comorbidity, most commonly cardiovascular, nervous system, and gastrointestinal conditions. Overall comorbidity burden was not associated with stage at diagnosis, although laxative-related prescribing was associated with later-stage disease. Increasing comorbidity burden reduced the likelihood of receiving any treatment and surgery, while associations varied across system-specific comorbidities. Higher comorbidity burden was also associated with increased all-cause and colorectal cancer-specific mortality, particularly among patients with respiratory, nervous system, and haematological/nutritional conditions. Screening uptake was not associated with overall comorbidity burden but did differ by system-specific comorbidity.

Prescribing-based multimorbidity was highly prevalent and strongly associated with treatment patterns and mortality among patients with colorectal cancer. System-specific multimorbidity measures provided greater discrimination than overall morbidity counts, highlighting the importance of considering distinct multimorbidity profiles when assessing cancer pathways and designing targeted interventions for optimising treatment and survival.

## INTRODUCTION

Multimorbidity, the co-existence of two or more long-term conditions,^1^ is common and has been reported to be up to three times more prevalent among people with cancer than in the general population.^2^ Multimorbidity is associated with higher mortality rates in people with cancer, particularly for cancers with a more favourable prognosis, including colorectal cancer.^3,4^ In Scotland, multimorbidity is present in most people aged 65 years and older, and presents much earlier in people living in the most deprived areas.^5^ Despite improvements in recent years, cancer survival in Scotland remains lower than in many comparable European countries,^6^ with five-year survival of approximately 50% based on the most recent national data (2018–2022).^7^ Differences in levels of multimorbidity are one plausible explanation for international differences in cancer survival.^8^

Multimorbidity has the potential to impact outcomes across the cancer control continuum, from prevention to early diagnosis, treatment, and survival.^⁹^ There are several mechanisms through which this may occur. Timely cancer diagnosis is an international healthcare priority and remains a major challenge in primary care.^10^ Multimorbidity can affect cancer stage, but the relationship is complex and different for specific comorbidities.^11^ While the impact on stage is not clearly understood,^12^ evidence suggests that people with coexisting conditions may experience longer diagnostic intervals^13^ and present with more advanced-stage disease for certain cancers.^14^ Cancer symptoms may be attributed to pre-existing chronic conditions, potentially prolonging diagnostic intervals through competing clinical demands or alternative explanations for symptoms.^13,15^ In primary care, where undifferentiated symptoms are managed alongside existing chronic disease, these mechanisms may be particularly relevant. Conversely, some studies suggest that multimorbidity may facilitate earlier diagnosis through increased healthcare contact and surveillance, although findings appear to vary by cancer type and healthcare setting.^16,17^

Among patients with colorectal cancer, comorbidity has been associated with longer times to surgery compared with patients without comorbid conditions.^3^ Older adults with cancer are also more likely to receive less intensive treatment, partly due to the presence of comorbidity, which may contribute to poorer clinical outcomes.^18–20^

Colorectal cancer is the fourth most common cancer in Scotland accounting for 12% (4,337 cases) of all cancer diagnoses in 2022.^7,21^ Common comorbidities among patients with cancer include congestive heart failure, diabetes mellitus, and chronic obstructive pulmonary disease, all of which have been associated with poorer overall survival.^19^ Utilising the ability to link disparate Scottish health records through the individual-specific Community Health Index (CHI), this study aimed to explore patterns of common comorbidities among colorectal cancer patients and examine the association between multimorbidity and colorectal cancer diagnosis, stage at diagnosis and provision of treatment, and both all-cause and colorectal cancer-specific mortality. By using prescribing data to derive system-specific comorbidity profiles, this study aimed to move beyond simple comorbidity counts to better reflect clinically active disease burden. We also examined the association between multimorbidity and pre-diagnostic participation in the Scottish Colorectal Cancer Screening Programme among colorectal cancer patients of screening age. All analyses used data from the Scottish Cancer Registry, a high-quality national dataset with near-complete population coverage and robust validation procedures.^21,22^

## METHODS

A population-based retrospective cohort study was carried out using electronic data-linkage and reported according to The Reporting of Studies Conducted using Observational Routinely-collected Health Data (RECORD) Statement.^23^ Patients diagnosed with colorectal cancer in Scotland were identified through the Scottish Cancer Registry. Data-linkage was carried out by the electronic Data Research and Innovation Services (eDRIS), Information Services Division (ISD) using the using the CHI number. Anonymised individual-level data was held within the NHS National Services Scotland (NSS) National Safe Haven and accessed by the researchers through a secure login that provided remote access via a virtual desktop. Requisite approvals were obtained from NHS Research Ethics and the Public Benefit and Privacy Panel (PBPP) for Health and Social Care.

### Data Sources

Linked datasets included: 1) Scottish Cancer Registry (Scottish Morbidity Records SMR06) which contains personal, demographic and clinical variables including diagnosis, staging and treatment information for all patients with a malignant diagnosis in Scotland; 2) Prescribing Information Systems (PIS) which contains information on all community dispensed prescriptions in Scotland; 3) General/acute inpatient and day case (SMR01) which has episode level data on hospital inpatient and day case discharges from hospitals in Scotland, including treatment information and operation procedure codes; 4) General Register Office (GRO) Death Register which provides information on cause of death; and 5) Scottish Bowel Screening Programme (SBoSP) contains personal and demographic information including data on participation and non-participation in the colorectal cancer screening programme.

### Study Cohort

Patients aged 18 years and older who were diagnosed with colorectal cancer in Scotland between 1 January 2010 and 31 December 2014 were identified using the Scottish Cancer Registry (SMR06).

### Variables

#### Exposures

Analyses explored associations between multimorbidity at the time of cancer diagnosis (using prescribing information as a proxy for multimorbidity - described below) and stage at diagnosis, cancer treatment and mortality. Stage at diagnosis was coded in accordance with the Dukes’ staging system, due to a significant amount of missing data for T (tumour), N (nodes) and M (metastasis) stage. Types of treatment included; surgery, radiotherapy and chemotherapy (including biological therapies). Survival analyses explored mortality from any-cause and colorectal cancer-specific mortality (based on the underlying International Classification of Diseases (ICD) code). Colorectal cancer screening data was available for those of screening age (50-74 years) living in Scotland between the years 2010 and 2014. Among those with screening data available, pre-diagnostic screening behaviour (participation in at least one screening round versus non-participation) was examined by logistic regression.

#### Covariates

The following covariates were adjusted for in each of the analyses; age, sex, deprivation using the Scottish Index of Multiple Deprivation (SIMD),^24^ rurality, method of presentation (clinical/screening/incidental/interval cancer/other), year of cancer incidence, and cancer site (Distal/Overlapping/Proximal/Rectum). In addition, stage at diagnosis (Dukes’ staging) was included in the cancer treatment and survival analyses, and cancer treatment (radiotherapy/chemotherapy/surgery/other) were included in the survival analyses.

#### Outcomes

##### Multimorbidity before diagnosis

Dispensed prescribing information from PIS was used as a proxy measure for calculating the presence of multimorbidity before diagnosis using the following definitions: 1) comorbidity, and 2) comorbidity by bodily system:

##### 1) Comorbidity (overall count)

Using the British National Formulary (BNF) classification system, comorbidity was derived by counting the number of unique prescribing subsections from which medications were dispensed. This approach was intended to capture active, treated morbidity, more closely reflecting clinical workload in primary care. Multiple prescriptions within the same BNF subsection (e.g. 040701) were counted once, representing a single comorbidity category.^25^ Long-term/chronic comorbidities were identified using regular prescriptions defined as being when patients had a dispensed prescription from the same BNF subsection within the first 84 days AND in the 85-365 days, preceding their cancer diagnosis.

##### 2) Comorbidity by bodily system

Comorbidity by bodily system (BNF Chapter) was calculated by counting the number of unique subsections from which a prescribing record was dispensed, within each BNF chapter. For regression analyses, comorbidity within each bodily system was categorised as 0, 1, or ≥2 conditions.

BNF codes relating to devices, non-chronic conditions or acute treatments were removed from the count: devices (peak flow meters, inhalers, diabetes monitoring), bowel cleaning preparations, stoma care, vaccines, contraceptives and emergency/acute treatments. Exclusions were based on expert consensus within the study team.

### Statistical Analyses

The prescribing dataset was explored using descriptive statistics including frequency counts and percentage of overall prescribing records based on the above-described comorbidity definitions. Frequencies of BNF chapters, sections and subsections were ranked to identify the most common comorbidities present at time of cancer diagnosis. In addition, the gastro-intestinal system (BNF chapter 1) was explored separately to identify concurrent gastrointestinal (GI) morbidities associated with a colorectal cancer diagnosis.

Data were initially explored by cross-tabulation and chi-squared test, for all demographic and clinical variables, by comorbidity count. Analyses of stage at diagnosis, treatment and previous screening participation, were by logistic regression methods, with stage at diagnosis reduced to early (Dukes’ A/B) and late (Dukes’ C/ D) cancer stage. Treatment analyses compared ‘no treatment’ and ‘at least one treatment’ including radiotherapy, chemotherapy and/or surgery, and ‘surgery’ compared to ‘no surgery’ as the main treatment modality for colorectal cancer. Survival analyses were by Kaplan-Meier (Log Rank test) and Cox Proportional Hazards regression. Univariate regression analyses explored associations between all demographic and clinical variables and comorbidity count (number of unique BNF subsections), and comorbidity by bodily system (BNF chapters).

Due to the large sample size, observed significant differences were highly significant, thus no official multiple testing adjustment was produced. A significance level of 5% was sufficient in the univariate analyses to allow variables to be included in the multivariable analyses. A significance level of less than or equal to 1% was required for variables to remain in the final models. All reported results are from the final multivariable models.

### Missing Data

Missing data by study exposure and outcome variables was explored using cross-tabulation and chi-squared test. Where missing values exceeded 10% (which was true for Dukes’ stage) a logistic regression was conducted to determine if comorbidity was an independent predictor of the absence or presence of staging data (1 or 0 respectively), with demographic and clinical variables added to the model. Missing Duke’s stage was calculated for cases with complete TNM stage data available (89 cases). A sensitivity logistic regression to predict cancer stage was conducted with the inclusion of all missing stage cases, where missing cases were treated as late-stage (Dukes’ C/D).

## RESULTS

### Cohort

A total of 19,043 patients were diagnosed with colorectal cancer [10,466 (55.0%) men and 8577 (45.0%) women] in Scotland during the five-year study period. 8639 (45.4%) patients died, of whom 7334 (85.5%) had cancer recorded as the underlying cause, 3943 (83.9%) for men and 3391 (86.1%) for women. Colorectal cancer was more common in people aged between 50 and 85 years, people living in urban areas, with broadly similar percentages of diagnoses across SIMD quintiles, for both sexes. Clinical presentation was the most common mode of detection, 78.1% for men and 83.1% for women, followed by screening, 19.1% for men and 13.7% for women. Study demographic and clinical variables stratified by sex are presented in Table 1.

**Table 1:** Study demographic and clinical variables stratified by sex.

| **N (%)** | **Male** | **Female** |
| --- | --- | --- |
| **All patients** | 10466 (55.0) | 8577 (45.0) |
| **Died** | 4700 (44.9) | 3939 (45.9) |
| **Cancer Death (ICD 10 C000-D499)**<br>Underlying cause | 3943 (83.9) | 3391 (86.1) |
| **Age group (at first SMR06)** |  |  |
| <50 | 491 (4.7) | 509 (5.9) |
| 50-64 | 2641 (25.2) | 1849 (21.6) |
| 65-74 | 3480 (33.3) | 2343 (27.3) |
| 75-84 | 2941 (28.1) | 2566 (29.9) |
| ≥85 | 913(8.7) | 1310 (15.3) |
| **Year of incidence** |  |  |
| 2010 | 2164 (20.7) | 1770 (20.6) |
| 2011 | 2190 (20.9) | 1749 (20.4) |
| 2012 | 2061 (19.7) | 1742 (20.3) |
| 2013 | 2043 (19.5) | 1696 (19.8) |
| 2014 | 2008 (19.2) | 1620 (18.9) |
| **SIMD quintile at time of incidence** |  |  |
| 1 - Most deprived | 2062 (19.7) | 1590 (18.5) |
| 2 | 2135 (20.4) | 1849 (21.6) |
| 3 | 2145 (20.5) | 1723 (20.1) |
| 4 | 2162 (20.7) | 1753 (20.4) |
| 5 – Least deprived | 1962 (18.7) | 1662 (19.4) |
| **Urban/rural (missing=8)** | 10459 | 8576 |
| 1 – Large urban area | 3622 (34.6) | 3013 (35.1) |
| 2 – Other urban area | 3243 (31.0) | 2701 (31.5) |
| 3 – Accessible small town | 924 (8.8) | 830 (9.7) |
| 4 – Remote small town | 504 (4.8) | 352 (4.1) |
| 5 – Accessible rural area | 1369 (13.1) | 1000 (11.7) |
| 6 – Remote rural area | 797 (7.6) | 680 (7.9) |
| **Presentation** |  |  |
| Screening | 1995 (19.1) | 1177 (13.7) |
| Incidental | 254 (2.4) | 231 (2.7) |
| Clinical presentation | 8173 (78.1) | 7128 (83.1) |
| Other (inc. Interval) | 22 (0.2) | 18 (0.2) |
| Not known | 22 (0.2) | 23 (0.3) |
| **Dukes (missing=2776)** | 9025 | 7242 |
| A | 1912 (21.2) | 1344 (18.6) |
| B | 2518 (27.9) | 2162 (29.9) |
| C1 | 1796 (19.9) | 1517 (20.9) |
| C2 | 304 (3.4) | 281 (3.9) |
| C | 292 (3.2) | 212 (2.9) |
| D | 2203 (24.4) | 1726 (23.8) |
| **Cancer Site** |  |  |
| Distal | 3127 (29.9) | 2211 (25.8) |
| Overlapping | 452 (4.3) | 517 (6.0) |
| Proximal | 3185 (30.4) | 3559 (41.5) |
| Rectum | 3702 (35.4) | 2290 (26.7) |
| **Cancer Treatment** |  |  |
| Surgery (missing=36) | 7798/10444 (74.7) | 6335/8563 (74.0) |
| Radiotherapy (missing=35) | 1456/10444 (13.9) | 830/8564 (9.7) |
| Chemotherapy (missing=79) | 3289/10419 (31.6) | 2517/8545 (29.5) |
| Other (missing =36) | 281/10445 (2.7) | 203/8562 (2.4) |
| No treatment (missing=46) | 1731/10439 (16.6) | 1641/8558 (19.2) |

ICD - International Classification of Diseases; SMR06 – Scottish Morbidity Records 06 (cancer registry); SIMD – Scottish Index of Multiple Deprivation

### Comorbidity

Most patients diagnosed with colorectal cancer had at least one comorbidity at the time of their cancer diagnosis (87%), 2482 (13%) were not prescribed and medicines before diagnosis [1504 men (14.4%) and 978 women (11.4%)]. Patients who died had a higher comorbidity count compared to those who were still alive at the end of the study period; median and interquartile range 4 (6) and 3 (5), respectively. All chi-squared tests showed statistically significant differences when comparing demographic and clinical variables by comorbidity count (Table 2)

**Table 2:**
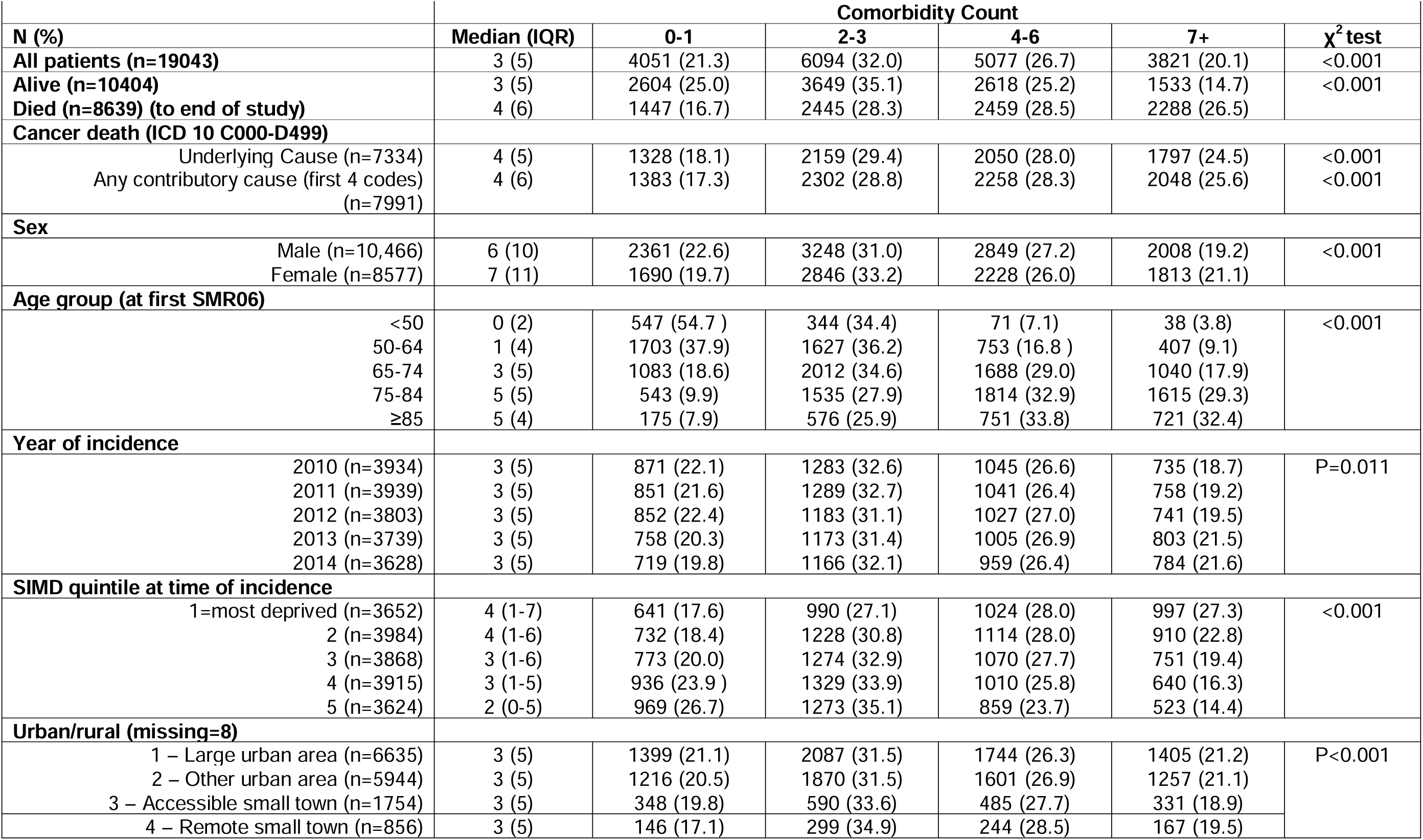
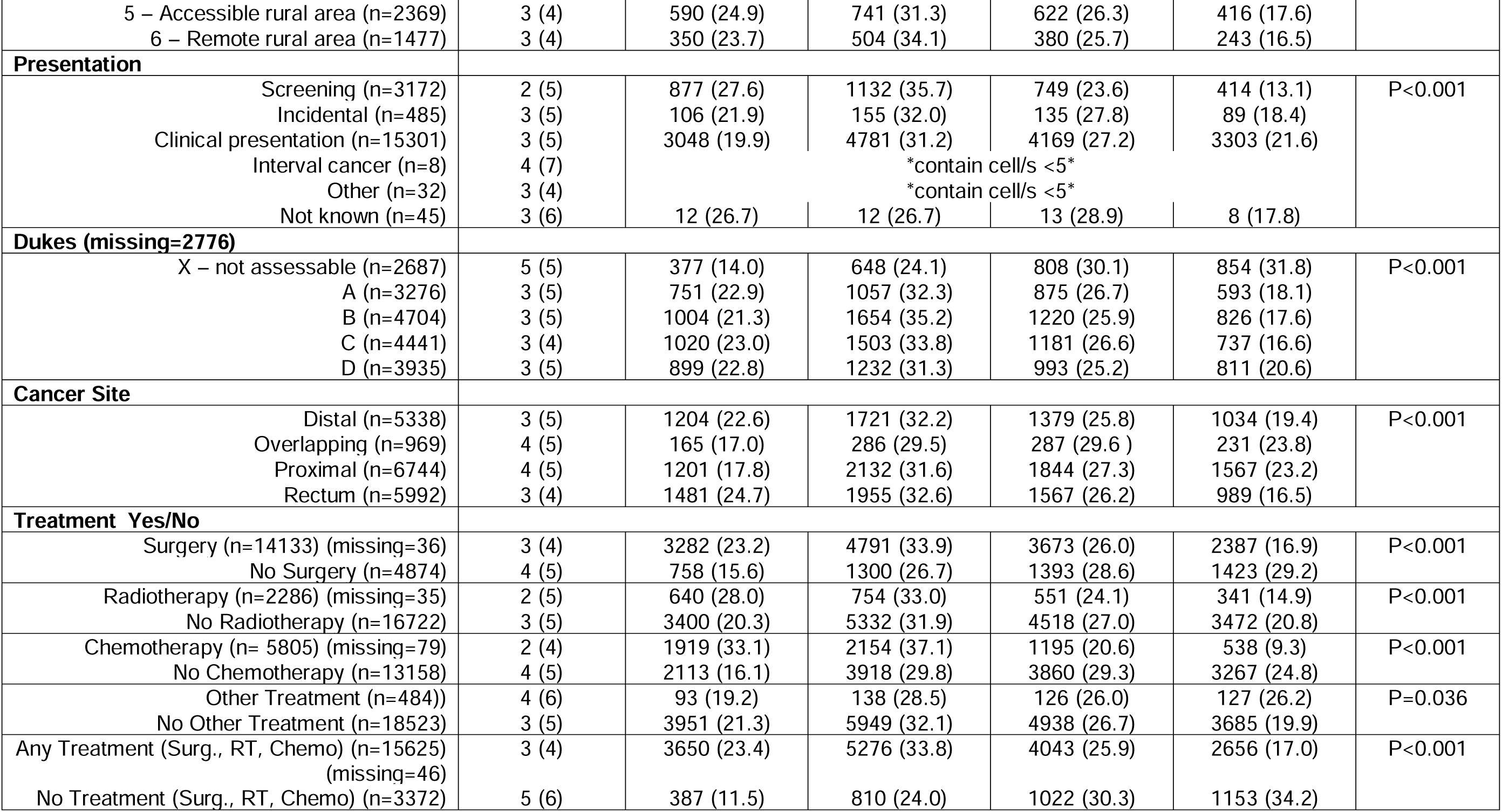
Demographic and clinical variables stratified by comorbidity count (defined by number of unique BNF subsections).

### Comorbidity by BNF chapter/subsection

The most common comorbidities, based on the total number of dispensed prescriptions by BNF chapter, were; cardiovascular (45.8%), the nervous system (14.9%) and the gastro-intestinal system (12.8%), for both sexes. By BNF section, the most prevalent comorbidities were conditions associated with lipid regulating drugs (8.7%), hypertension and heart failure (8.5%), and analgesics (8.3%), and by BNF subsection, lipid regulating drugs (8.7%), renin-angiotensin system drugs (7.6%) and proton pump inhibitors (6.5%).

Within the gastro-intestinal system (BNF chapter 1), comorbidity associated with the following sections were most common; antisecretory drugs and mucosal protectants, laxatives, dyspepsia and gastro-oesophageal reflux, and antispasmodics and other drugs altering gut motility.

### Cancer Stage

Staging data information was missing for 2687 (14%) patients. Missing data for Dukes’ stage differed significantly across comorbidity count with an increasing trend towards missing data with increasing comorbidity [X^2^ test: p<0.001, test for trend p<0.001]. A sensitivity analysis by logistic regression was conducted to predict missing stage data [missing (1) or not missing (0)], all demographic and clinical variables at the univariate level were significant predictors of missing Dukes’ stage. At the multivariable level all variables remained significant, except sex. Comorbidity count predicted missing Dukes’ stage [Adjusted OR 1.08 for each additional morbidity, 95% CIs (1.07, 1.09), p<0.001] (Supplementary Table S1).

Examining only the patients with staging data (86%), the multivariable model found that younger age, overlapping tumour site and clinical presentation were all statistically significant predictors of late-stage cancer (Dukes’ C/D). Comorbidity count was not associated with late-stage colorectal cancer at the univariate or multivariable level [Adjusted OR 0.998, 95% CIs (0.99, 1.01), p=0.67] (Table 3). In the model considering comorbidity by bodily system, only prescribing for laxatives (BNF section 106) was associated with an increased risk of late-stage cancer with risk increasing with prescribing from multiple subsections [Adjusted OR 1.18, 95% CIs (1.04, 1.34), p=0.01; Adjusted OR 1.54 95% CIs 1.54 (1.10, 2.17), P=0.01], for prescribing from 1 and 2+ subsections respectively (Table 4).

**Table 3:**
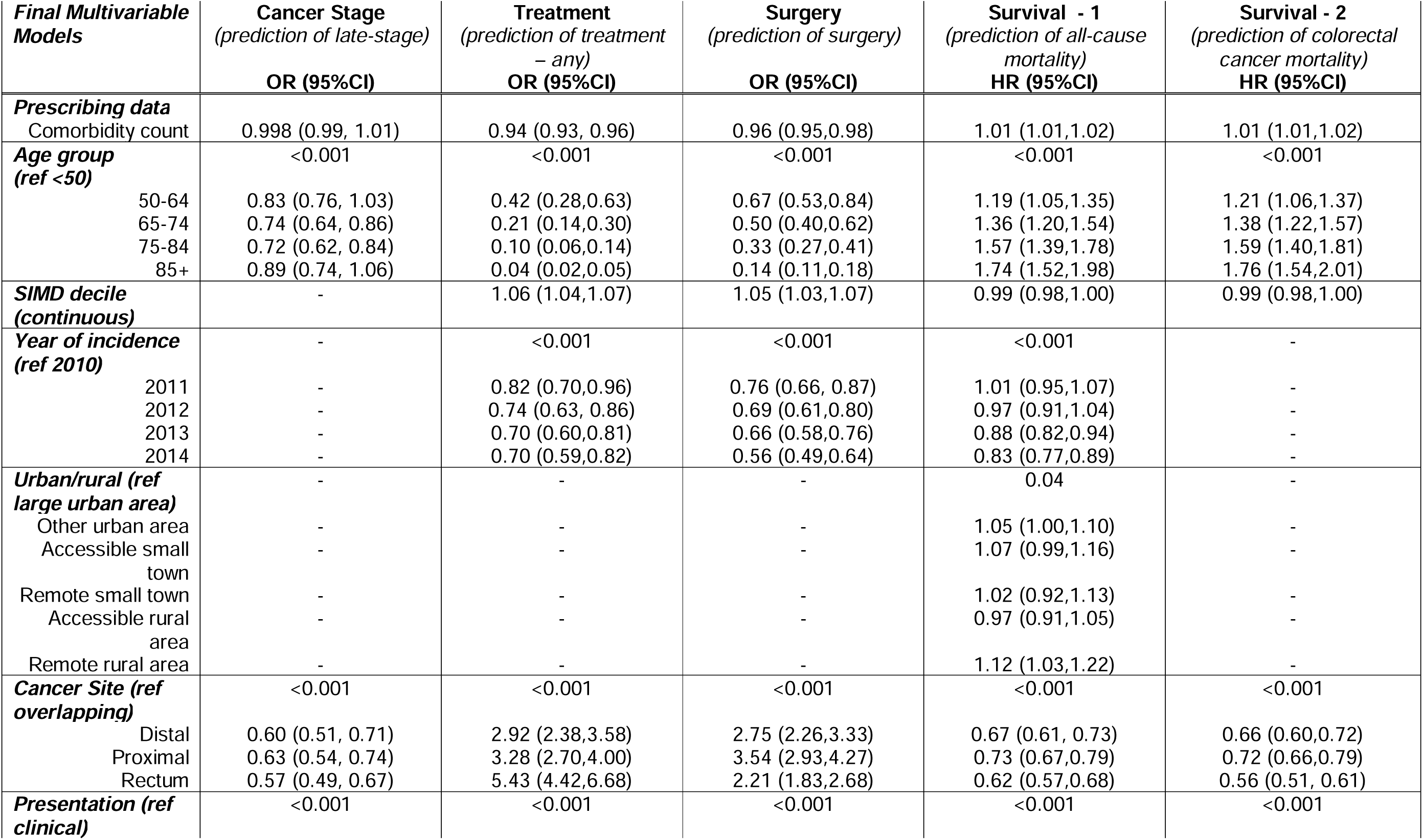
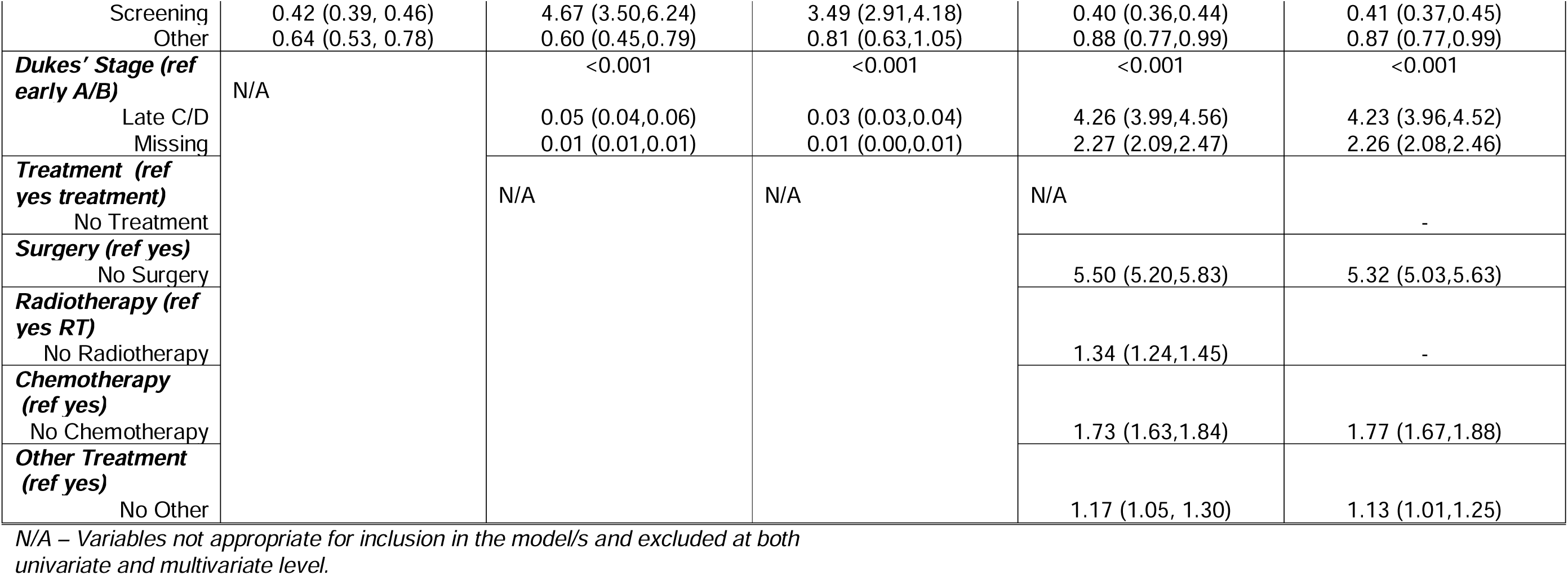
Multivariable regression for cancer stage, treatment and mortality and comorbidity (defined by number of unique BNF subsections).

**Table 4:** Multivariable regression models for cancer stage, treatment and mortality and comorbidity by bodily system (BNF Chapters)

| Final Multivariable Models | Cancer Stage Ø<br>(prediction of late-stage) | Treatment ¥<br>(prediction of treatment – any) | Surgery β<br>(prediction of surgery) | Survival 1 #<br>(prediction of all-cause mortality) | Survival 2 μ<br>(prediction of colorectal cancer mortality) |
| --- | --- | --- | --- | --- | --- |
| BNF Chapters | OR (95%CI) | OR (95%CI) | OR (95%CI) | HR (95%CI) | HR (95%CI) |
| 2. Cardiovascular |  | 0.01 | - | - | - |
| 1 | - | 1.30 (1.10,1.54) | - | - | - |
| 2+ | - | 1.06 (0.94,1.19) | - | - | - |
| 3. Respiratory system |  |  |  | 0.02 | 0.02 |
| 1 | - | - | - | 1.01 (0.93,1.10) | 1.01 (0.92,1.09) |
| 2+ | - | - | - | 1.12 (1.03,1.21) | 1.12 (1.04,1.21) |
| 4. Nervous system |  | <0.001 | <0.001 | 0.01 | 0.02 |
| 1 | - | 0.76 (0.67,0.85) | 0.83 (0.74,0.92) | 1.05 (0.99,1.10) | 1.05 (0.99,1.10) |
| 2+ | - | 0.61 (0.53,0.71) | 0.71 (0.62,0.81) | 1.10 (1.03,1.17) | 1.09 (1.02,1.16) |
| 6. Endocrine system |  | 0.01 | 0.03 |  |  |
| 1 | - | 1.04 (0.92,1.18) | 1.09 (0.97,1.22) | - | - |
| 2+ | - | 0.72 (0.58,0.90) | 0.81 (0.66,0.996) | - | - |
| 7. Genito-urinary system |  | 0.01 |  |  |  |
| 1 | - | 1.31 (1.09,1.56) | - | - | - |
| 2+ | - | 0.89 (0.46,1.71) | - | - | - |
| 8. Malignant disease |  |  |  | <0.001 |  |
| 1 | - | - | - | 1.28 (1.11,1.47), 0.001 | - |
| 2+ | - | - | - | 2.18 (1.20, 3.94), 0.01 | - |
| 9. Blood and nutrition |  | <0.001 | <0.001 | <0.001 | <0.001 |
| 1 | - | 0.72 (0.63,0.820) | 0.88 (0.77,0.997) | 1.05 (0.99,1.12) | 1.05 (0.99,1.11) |
| 2+ | - | 0.49 (0.39,0.61) | 0.61 (0.49,0.77) | 1.26 (1.14,1.39) | 1.26 (1.14,1.39) |
| **BNF Sections** |  |  |  |  |  |
| Gastro-intestinal system only (Ch.1) |  |  |  |  |  |
| 106 Laxatives | 0.002 | Not included in models |  |  |  |
| 1 | 1.18 (1.04,1.34) |  |  |  |  |
| 2+ | 1.54 (1.10,2.17) |  |  |  |  |

Ø - Cancer Stage model adjusted for: age group, cancer site and presentation.
¥ - Treatment model adjusted for: sex, age group, SIMD, year of incidence, cancer site, presentation and cancer stage.
β - Surgery model adjusted for: age group, SIMD, year of incidence, cancer site, presentation and cancer stage.

### Sensitivity analysis

The above model was repeated, recoding all missing cancer stage cases as late stage (worst case scenario). This model showed a statistically significant association between increasing comorbidity count and late-stage cancer [Adjusted OR 1.02, 95% CIs (1.01, 1.02), p=0.003]. As before younger age, overlapping cancer site and clinical presentation were significant predictors of late cancer stage. However, with missing cases included SIMD was also now a significant predictor of late-stage cancer [adjusted OR 0.98, 95% CIs (0.97, 0.99), p<0.001)]. The final model exploring comorbidity by bodily system (including BNF chapters and sections - chapter one only) found comorbidities associated with the nervous system (chapter 4), blood and nutrition (chapter 9) and laxatives (subsection 106) to be statistically significant predictors of late-stage cancer.

### Treatment

Logistic regression to predict missing treatment information was not required due to the small numbers of records missing this variable (0.2%) (Table 2). Surgery was the most common treatment modality (74.2%), followed by chemotherapy (30.5%), and radiotherapy (12.0%). The treatment outcome was defined as the presence of ‘at least one’ of surgery, chemotherapy or radiotherapy, compared to no treatment. In the final multivariable model, male sex, younger age, lower levels of deprivation, earlier year of diagnosis, non-overlapping tumour sites, screen-detected cancer, and earlier-stage disease were all associated with a higher likelihood of receiving at least one treatment modality. Increasing comorbidity burden was associated with a statistically significant reduction in the likelihood of receiving at least one treatment modality [Adjusted OR 0.94, 95% CIs (0.93, 0.96), p<0.001] (Table 3) consistent with treatment decisions balancing benefit against comorbid burden and frailty. In the multivariable model for comorbidity by bodily system; the nervous system, the endocrine system, and blood and nutrition were associated with a statistically significant reduction in the likelihood of receiving ‘at least one’ treatment. Conversely, comorbidities associated with the cardiovascular system and the genito-urinary system were statistically significant predicators of receiving ‘at least one’ treatment (Table 4).

### Surgery

In the final multivariable model, younger age, lower levels of deprivation, earlier year of diagnosis, early-stage disease (Dukes’ A/B), non-overlapping tumour sites, and screen-detected cancers were all statistically significant predictors of receiving surgery (Table 3). Increasing comorbidity burden was associated with a statistically significant reduction in the likelihood of receiving surgery [Adjusted OR 0.96, 95% CIs (0.95,0.98), p<0.001] (Table 3). Comorbidities affecting the nervous system, as well as endocrine disorders (≥2 conditions) and blood and nutritional conditions (≥2 conditions), were associated with a reduced probability of receiving surgery (Table 4).

### All-cause mortality

Univariately, increasing comorbidity count was associated with an increased risk of all-cause mortality. Results from the multivariable Cox proportional hazards regression analysis showed that older age, higher levels of deprivation, earlier year of diagnosis, clinically detected tumours, late-stage or stage-unknown cancers, overlapping tumour sites, and rural residence were all statistically significant predictors of all-cause mortality. Receipt of treatment, whether it be surgery (strongest association), chemotherapy, radiotherapy, or other was associated with a reduced risk of all-cause mortality (Table 3). A statistically significant positive association was observed between comorbidity burden and all-cause mortality, with increasing comorbidity count associated with a greater risk of death during the follow-up period (up to five years) [adjusted HR=1.01 (95% CI 1.01, 1.02)]. Comorbidities associated with the following bodily systems; respiratory, nervous system, malignant disease, and blood and nutrition were associated with an increased risk of all-cause mortality during the study follow up (up to five years) (Table 4).

### Colorectal cancer-specific mortality

Of the 8639 deaths, 7334 (84.9%) were attributable to colorectal cancer as the underlying/primary cause of death (Table1). Increasing comorbidity burden was associated with a higher risk of colorectal cancer-specific mortality during follow-up (up to five years) (adjusted HR = 1.01, 95% CI 1.01–1.02; p < 0.001) (Table 2). A positive association was observed for comorbidities of the respiratory and nervous systems, as well as blood and nutritional conditions, where higher levels of comorbidity (generally defined as prescribing from two or more subsections) were associated with an increased risk of colorectal cancer-specific mortality during follow-up (Table 4).

For all regression models, comorbidity by bodily systems provided a better description of the data compared to comorbidity (overall count) (p<0.001), despite loss of degrees of freedom. The final models for comorbidity by bodily systems, including demographic and clinical variables are provided as Supplementary Table S2.

### Colorectal cancer screening

Among patients aged 50–84 years with available screening data (n = 9,961), 5,885 (59.1%) had participated in the colorectal screening programme at least once prior to diagnosis (57.1% of men and 61.9% of women). A higher proportion of deaths occurred among non-responders (those who had not participated in screening) compared with responders (58.2% vs 41.8%). In multivariable analyses, female sex, older age (within screening eligibility limits), lower deprivation, more recent diagnosis year, and rural residence were associated with higher screening uptake. No significant association was observed between overall comorbidity count and screening participation [adjusted OR=0.99 (95% CI 0.98, 1.00)]. However, system-specific comorbidities showed variation, with cardiovascular, genitourinary, and musculoskeletal conditions significantly associated with increased screening uptake, while respiratory, nervous system, and blood and nutritional conditions were associated with reduced uptake (Table 5).

**Table 5:**
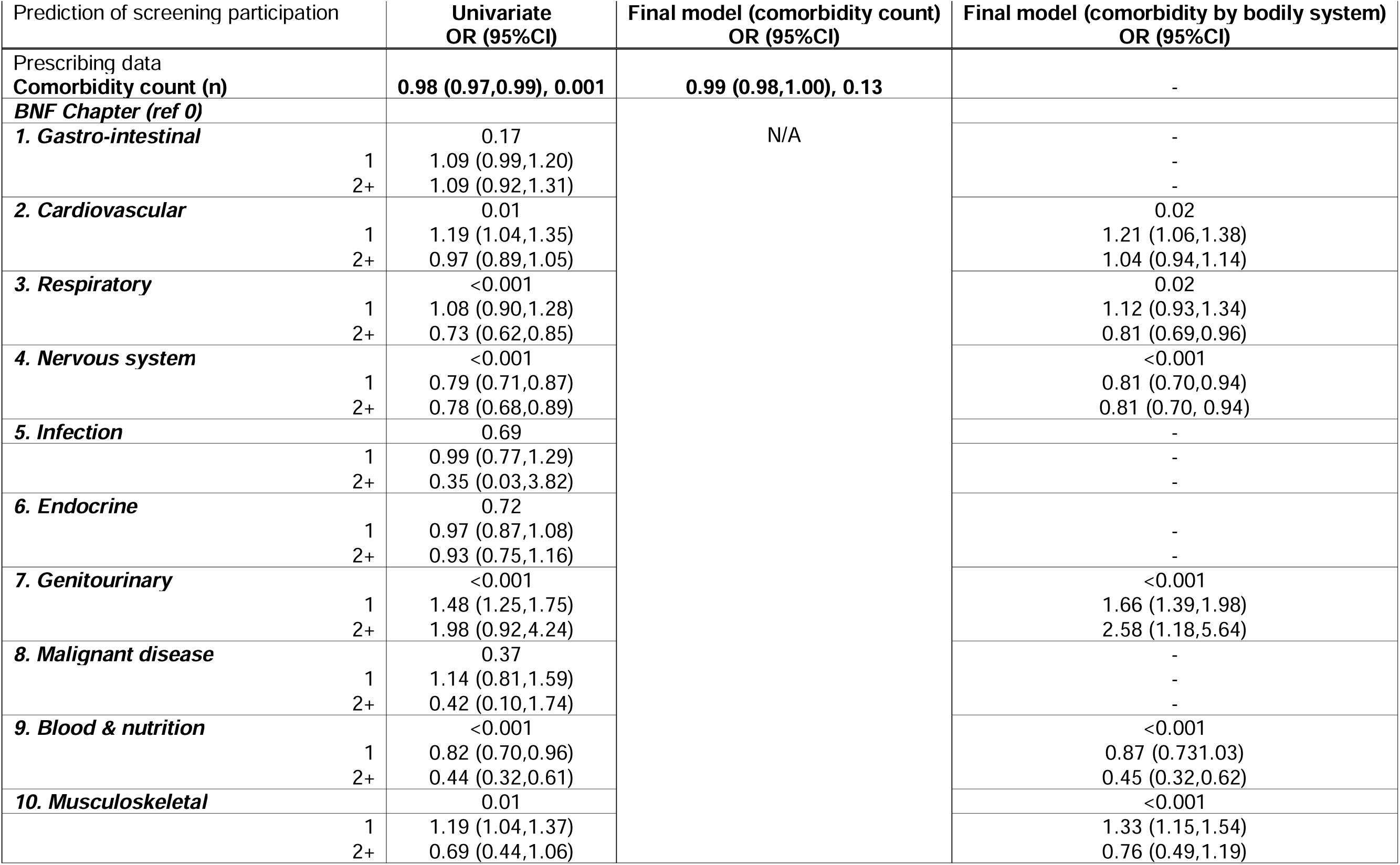
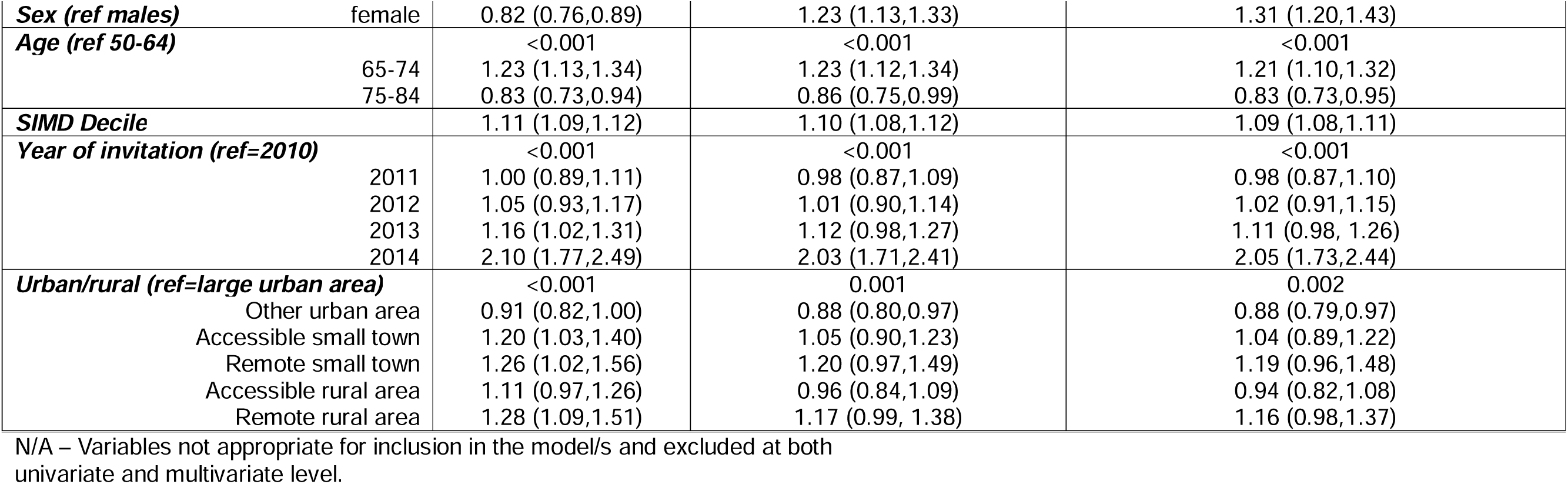
Univariate and multivariable regression models for colorectal cancer screening participation and comorbidity.

## DISCUSSION

### Summary Results

In this large, population-based study linking national cancer, prescribing, hospital, screening, and mortality datasets, prescribing-based multimorbidity was highly prevalent at the time of diagnosis with colorectal cancer in Scotland, with almost nine in ten patients having at least one comorbidity. Although the overall comorbidity count was not associated with stage at diagnosis, notably, laxative prescribing was the only system-specific factor associated with later-stage disease. Increasing overall comorbidity burden was strongly associated with reduced treatment uptake (any) and surgery, and with higher all-cause and colorectal cancer-specific mortality. However, system-specific multimorbidity profiles provided clearer insight into these variations, indicating that the type of comorbidity, rather than the number alone, is more clinically informative when assessing cancer pathways and outcomes. Importantly, the type of comorbidity appeared more clinically informative than the number alone

### Comparison with existing literature

Our results align with previous work showing that multimorbidity is common in cancer populations and associated with poorer survival.^3,4^ However, the wider evidence base remains heterogeneous, with studies employing varying definitions of multimorbidity and largely relying on diagnosis-based indices, which may under-represent active disease. Such methodological variation limits comparability and may obscure genuine associations between multimorbidity and diagnostic timeliness. The absence of an association between overall comorbidity count and stage at diagnosis is consistent with recent cohort studies.^26,27^ In contrast, the association between laxative-related prescribing and later-stage cancer may reflect diagnostic complexity in primary care, where symptoms such as constipation may be attributed to benign or chronic conditions before malignancy is considered potentially contributing to delayed investigation. Such mechanisms have been proposed in relation to diagnostic delay but remain underexplored in empirical research.^10,13^ Younger age remained independently associated with late-stage colorectal cancer despite adjustment for mode of detection, consistent with systematic review evidence showing that adults under 50 have higher odds of advanced-stage disease at diagnosis compared with older patients,^28^ underscoring that this disparity is not explained by screening or diagnostic pathway differences alone.

The association between multimorbidity and reduced receipt of treatment is well documented.^18,19^ In this study, system-specific comorbidities, particularly respiratory, neurological, and blood and nutrition conditions, were associated with higher mortality, while overall comorbidity count was a weaker predictor. These findings suggest that certain conditions may exert disproportionate effects through reduced physiological reserve, treatment contraindications, or competing mortality risks.^20,29–30^ In contrast, higher receipt of treatment among patients with cardiovascular and genito-urinary comorbidities may reflect greater healthcare contact or earlier clinical review. Overall, these results highlight limitations of aggregate comorbidity measures and support the value of system-specific profiling in understanding multimorbidity and cancer treatment interactions.

Screening participation was not associated with overall comorbidity burden, consistent with mixed and non-linear associations reported previously.^31,32^ However, system-specific comorbidities showed divergent associations with screening uptake, suggesting that factors such as functional status, symptom burden, and healthcare engagement vary across comorbidity profiles. This supports the argument that the influence of multimorbidity on screening behaviour depends on the nature and severity of underlying conditions rather than the number of comorbidities alone.^33^

### Strengths and limitations

The study benefited from the ability to use the Community Health Index (CHI) to link disparate datasets in Scotland to obtain individual-level data on cancer registration, prescribing records, treatment and operation procedures, bowel cancer screening participation, and mortality. Although the Scottish Cancer Registry provides a comprehensive dataset of cancer cases and staging, for the time period included in our study the level of missing stage data was 14% and was significantly associated with increasing comorbidity. However, among the 86% of cases with complete stage data, comorbidity count was not associated with stage at either univariate or multivariate level. We acknowledge the impact of missing data is not known. However, sensitivity analyses indicated that findings were robust. In a worst-case scenario analysis, recoding missing stage as late stage did not materially change results, and socioeconomic deprivation became a stronger predictor, suggesting potential underestimation in complete-case analyses.

The study also benefited from the novel use of community dispensed prescribing data to derive system-specific comorbidity profiles. Compared with diagnosis-based measures, prescribing-based approaches may better reflect active disease burden and healthcare use, and decision making in primary care. Prescribing measures also have their limitations, including potential misclassification due to non-adherence (although this study used dispensed prescribing data rather than issued prescriptions), incomplete capture of over-the-counter and hospital-prescribed medications, and limited information on disease severity. Beyond these methodological considerations, analysis of more recent diagnostic cohorts would be valuable to determine whether these patterns persist in the context of evolving cancer care pathways and changing population-level multimorbidity.

### Implications for research and/or practice

These findings underscore that multimorbidity is not a uniform exposure, and that system-specific profiling of comorbid conditions may better capture the heterogeneous ways in which coexisting disease influences the cancer pathway. Understanding how particular comorbidities influence diagnostic pathways, treatment decisions, and survival may support more personalised care, risk stratification, and prehabilitation - reinforcing diagnostic vigilance in patients with overlapping symptoms.

Future research should explore mechanisms linking specific comorbidities to poorer outcomes and assess whether system-specific measures of multimorbidity can improve early diagnosis and treatment optimisation.

Overall, these findings highlight multimorbidity as a key determinant of colorectal cancer treatment and outcomes, supporting a move beyond aggregate comorbidity measures in research and clinical care.

## Additional information

### Author contributions

KB was the lead author and wrote the first draft of the manuscript. KB and LW co-led the methodology, investigation, data curation, and formal analysis. KB, DW, SM, BG, HW, DB, GH, and CC contributed to conceptualisation and funding acquisition, with CC additionally providing supervision. All authors contributed to the analysis and interpretation, reviewed and edited the manuscript, and approved the final version for submission.

### Ethical approval

Favourable ethical opinion received from the Health Research Authority, East Midlands – Derby Research Ethics Committee on 16^th^ April 2016. REC reference 16/EM/0150.

## Acknowledgements

We thank the electronic Data Research and Innovation Service (eDRIS) and the Public Benefit and Privacy Panel for Health and Social Care (PBPP) at NHS National Services Scotland for support with approvals, data linkage and access to the data within the National Safe Haven. We also thank patient representatives from the SCAN Patient Involvement Group of the Southeast Scotland Cancer Network who provided detailed feedback on the initial research design.

## Data Availability Statement

The data used in this study are held by NHS National Services Scotland (NSS) within the National Safe Haven and are not publicly available due to patient confidentiality and data governance restrictions. The authors accessed the data under a time-limited approval granted by the Public Benefit and Privacy Panel for Health and Social Care (PBPP). In accordance with National Safe Haven governance arrangements, the authors no longer have direct access to the underlying data. Any further analyses would require a new application and approval process through the PBPP. Researchers wishing to access the data may apply via NHS Scotland Information Governance. The authors did not receive any special access privileges unavailable to other researchers.

## Supporting information captions

Table S1: Univariate and multivariable regression models to predict missing Dukes’ stage.

Table S2: Final multivariable regression models for treatment, surgery and all-cause and colorectal specific mortality by comorbidity by bodily system (BNF Chapters).

