## Supplementary Information for "System-specific multimorbidity derived from prescribing data predicts colorectal cancer outcomes: a Scottish data-linkage study"

**Colorectal Cancer Cohort**

**Table S1: Univariate and multivariable regression models to predict missing Dukes’ stage.**

| *Prediction of missing* | **Univariate** | **Multivariable (excluding comorbidity)** | **Final model (including comorbidity)** |
| --- | --- | --- | --- |
|  | **OR (95%CI)** | **OR (95%CI)** | **OR (95%CI)** |
| ***Prescribing data*** |  |  |  |
| Chronic count (n) | 1.12 (1.11, 1.14), <0.001 | - | 1.08 (1.07, 1.09), <0.001 |
| ***Sex (ref female)*** | 0.869 (0.80,0.94) | 0.97 (0.89, 1.07) | - |
| ***Age (ref <50)*** | <0.001 | <0.001 | <0.001 |
| 50-64 | 0.72 (0.56, 0.92) | 0.87 (0.67, 1.12) | 0.79 (0.61, 1.03) |
| 65-74 | 1.02 (0.80, 1.30) | 1.33 (1.04, 1.70) | 1.09 (0.85, 1.40) |
| 75-84 | 2.16 (1.71, 2.73) | 2.59 (2.03, 3.29) | 1.96 (1.53, 2.51) |
| 85+ | 7.28 (5.74, 9.24) | 8.88 (6.94, 11.36) | 6.70 (5.21, 8.61) |
| ***SIMD decile*** | 0.95 (0.94, 0.97) | 0.95 (0.93, 0.96) | 0.96 (0.95, 0.98) |
| ***Year of incidence (ref=2010)*** | <0.001 | <0.001 | <0.001 |
| 2011 | 0.74 (0.66, 0.84) | 0.72 (0.63, 0.82) | 0.71 (0.62, 0.81) |
| 2012 | 0.78 (0.69, 0.88) | 0.74 (0.65, 0.85) | 0.73 (0.64, 0.84) |
| 2013 | 0.73 (0.65, 0.83) | 0.69 (0.60, 0.79) | 0.67 (0.59, 0.77) |
| 2014 | 0.64 (0.56, 0.72) | 0.59 (0.51, 0.68) | 0.57 (0.50, 0.66) |
| ***Urban/rural (ref=large urban area)*** | 0.011 | 0.01 | 0.02 |
| Other urban area | 0.90 (0.82, 0.997) | 0.93 (0.84, 1.04) | 0.93 (0.84, 1.04) |
| Accessible small town | 0.83 (0.71, 0.96) | 0.85 (0.72, 1.00) | 0.84 (0.71, 0.99) |
| Remote small town | 0.79 (0.64, 0.98) | 0.78 (0.62, 0.98) | 0.78 (0.62, 0.98) |
| Accessible rural area | 0.88 (0.77, 1.00) | 0.99 (0.85, 1.14) | 0.99 (0.86, 1.15) |
| Remote rural area | 0.79 (0.67, 0.94) | 0.75 (0.63, 0.90) | 0.78 (0.65, 0.94) |
| ***Site (ref overlapping)*** | <0.001 | <0.001 | <0.001 |
| Distal | 0.41 (0.34, 0.49) | 0.54 (0.44, 0.65) | 0.54 (0.45, 0.66) |
| Proximal | 0.48 (0.40, 0.56) | 0.49 (0.41, 0.59) | 0.49 (0.41, 0.59) |
| Rectum | 0.77 (0.66, 0.91) | 1.08 (0.90, 1.29) | 1.13 (0.94, 1.35) |
| ***Method of detection (ref clinical)*** | <0.001 | <0.001 | <0.001 |
| Screening | 0.21 (0.18, 0.26) | 0.39 (0.32, 0.47) | 0.40 (0.33, 0.49) |
| Other | 1.36 (1.09, 1.69) | 1.69 (1.34, 2.13) | 1.72 (1.36, 2.17) |

**Table S2: Final multivariable regression models for treatment, surgery and all-cause and colorectal specific mortality**

**by comorbidity by bodily system (BNF Chapters).**

| **Comorbidity by bodily system (BNF chapter)** | | | | | |
| --- | --- | --- | --- | --- | --- |
|  | **Treatment**  *(prediction of treatment)* | **Surgery**  *(prediction of surgery)* | | **All-cause mortality**  *(prediction of death)* | **Colorectal cancer-specific mortality** *(prediction of death)* |
|  | **OR (95%CI)** | **OR (95%CI)** | | **HR (95%CI)** | **HR (95%CI)** |
| ***Sex (ref female)*** | 1.24 (1.12,1.38) | - | | - | - |
| ***Age (ref <50)*** | <0.001 | <0.001 | | <0.001 | <0.001 |
| 50-64 | 0.40 (0.27,0.60) | 0.65 (0.53,0.81) | | 1.20 (1.06,1.36) | 1.21 (1.07,1.38) |
| 65-74 | 0.19 (0.13,0.28) | 0.47 (0.38,0.59) | | 1.38 (1.22,1.56) | 1.40 (1.23,1.58) |
| 75-84 | 0.09 (0.06,0.13) | 0.31 (0.25,0.38) | | 1.60 (1.41,1.81) | 1.61 (1.42,1.83) |
| 85+ | 0.03 (0.02,0.05) | 0.14 (0.11,0.17) | | 1.75 (1.53,2.00) | 1.78 (1.56,2.03) |
| ***SIMD Decile*** | 1.05 (1.03,1.07) | 1.05 (1.03,1.06) | | 0.99 (0.98,1.00) | 0.99 (0.98,1.00) |
| ***Year of incidence (ref=2010)*** | <0.001 | <0.001 | | <0.001 | - |
| 2011 | 0.83 (0.71,0.97) | 0.76 (0.67,0.87) | | 1.00 (0.94,1.07) | - |
| 2012 | 0.74 (0.63,0.86) | 0.70 (0.61,0.80) | | 0.97 (0.91,1.04) | - |
| 2013 | 0.70 (0.59,0.81) | 0.66 (0.58,0.76) | | 0.88 (0.82,0.94) | - |
| 2014 | 0.71 (0.61,0.84) | 0.57 (0.50,0.65) | | 0.82 (0.76,0.88) | - |
| ***Urban/rural***  ***(ref=large urban)*** | - | - | | 0.03 | - |
| Other urban area | - | - | | 1.05 (1.00,1.11) | - |
| Accessible small town | - | - | | 1.07 (0.99,1.16) | - |
| Remote small town | - | - | | 1.02 (0.92,1.14) | - |
| Accessible rural area | - | - | | 0.98 (0.91,1.06) | - |
| Remote rural area | - | - | | 1.13 (1.04,1.23) | - |
| ***Site (ref overlapping)*** | <0.001 | <0.001 | | <0.001 | <0.001 |
| Distal | 2.88 (2.35,3.54) | 2.69 (2.22,3.26) | | 0.67 (0.61,0.73) | 0.66 (0.61,0.72) |
| Proximal | 3.35 (2.74,4.08) | 3.53 (2.92,4.27) | | 0.72 (0.66,0.79) | 0.72 (0.66,0.78) |
| Rectum | 5.41 (4.39,6.66) | 2.17 (1.79,2.62) | | 0.62 (0.57,0.68) | 0.56 (0.51,0.61) |
| ***Method of Detection (ref clinical)*** | <0.001 | <0.001 | | <0.001 | <0.001 |
| Screening | 4.52 (3.38,6.04) | 3.44 (2.87,4.13) | | 0.40 (0.37,0.45) | 0.41 (0.37,0.45) |
| Other | 0.59 (0.45,0.78) | 0.81 (0.63,1.06) | | 0.88 (0.77,0.99) | 0.88 (0.77,0.99) |
| ***Dukes Stage (ref early)***  Late (C&D)  Missing stage | <0.001  0.05 (0.04,0.06)  0.01 (0.01,0.01) | <0.001  0.03 (0.03,0.04)  0.01 (0.004,0.01) | | <0.001  4.26 (3.99,4.56)  2.25 (2.07,2.44) | <0.001  4.23 (3.96,4.52)  2.24 (2.07,2.44) |
| ***Treatment (ref yes)*** | Not included | Not included | | Not included | Not included |
| ***Surgery (ref yes)*** | - | - | | 5.49 (5.19,5.82) | 5.29 (5.00,5.60) |
| ***Chemotherapy (ref yes)*** | - | - | | 1.72 (1.62,1.82), | 1.76 (1.66,1.87) |
| ***Radiotherapy (ref yes)*** | - | - | | 1.34 (1.24,1.44) | - |
| ***Other (ref yes)*** | - | - | | 1.18 (1.06,1.31) | 1.14 (1.02,1.26) |
| ***BNF Chapter (ref 0)*** | | |  | | |
| ***1. Gastro-intestinal*** |  |  | |  |  |
| 1 | - | *-* | | - | *-* |
| 2+ | - | *-* | | - | *-* |
| ***2. Cardiovascular*** | 0.01 |  | |  |  |
| 1 | 1.30 (1.10,1.54) | *-* | | - | - |
| 2+ | 1.06 (0.94,1.19) | *-* | | - | - |
| ***3. Respiratory*** |  |  | | 0.02 | 0.02 |
| 1 | - | *-* | | 1.01 (0.93,1.10) | 1.01 (0.92,1.09), 0.89 |
| 2+ | - | *-* | | 1.12 (1.03,1.21) | 1.12 (1.04,1.21), 0.01 |
| ***4. Nervous system*** | <0.001 | <0.001 | | 0.01 | 0.02 |
| 1 | 0.76 (0.67,0.85) | 0.83 (0.74,0.92) | | 1.05 (0.99,1.10) | 1.05 (0.99,1.10), 0.09 |
| 2+ | 0.61 (0.53,0.71) | 0.71 (0.62,0.81) | | 1.10 (1.03,1.17) | 1.09 (1.02,1.16), 0.01 |
| ***5. Infection*** |  |  | |  |  |
| 1 | - | *-* | | - | *-* |
| 2+ | - | *-* | | - | *-* |
| ***6. Endocrine*** | 0.01 | 0.03 | |  |  |
| 1 | 1.04 (0.92,1.18) | 1.09 (0.97,1.22) | | *-* | *-* |
| 2+ | 0.72 90.58,0.90) | 0.81 (0.66,0.99) | | *-* | *-* |
| ***7. Genitourinary***  1  2+ | 0.01  1.31 (1.09,1.56)  0.89 (0.46,1.71) | *-*  *-* | | *-*  *-* | *-*  *-* |
| ***8. Malignant disease*** |  |  | | <0.001 | - |
| 1 | - | - | | 1.28 (1.11,1.47) | - |
| 2+ | - | - | | 2.18 (1.20, 3.94) | - |
| ***9. Blood & nutrition*** | <0.001 | <0.001 | | <0.001 | <0.001 |
| 1 | 0.72 (0.63,0.82) | 0.88 (0.77,0.99) | | 1.05 (0.99,1.12) | 1.05 (0.99,1.11), 0.12 |
| 2+ | 0.49 (0.39,0.61) | 0.61 (0.49,0.77) | | 1.26 (1.14,1.39) | 1.26 (1.14,1.39), <0.001 |
| ***10. Musculoskeletal*** |  |  | |  |  |
| 1 | - | - | | - | *-* |
| 2+ | - | - | | - | *-* |
